# Admission and discharge profiles of people with MS accessing in-patient rehabilitation in Canada

**DOI:** 10.1101/2023.06.25.23291871

**Authors:** Kedar K. V. Mate, Nikki Ow, Stanley Hum, Nancy Mayo

## Abstract

**Background:** Rehabilitation is the mainstay of management for people with disabilities of neurological origin to maximize function and reduce disability. Access to in-patient rehabilitation is usually reserved for people after crises or those who are discharged from acute care such as in stroke or trauma. Access to people with Multiple Sclerosis (MS) differs across countries and unknown for Canada. With the progression of MS, quality of life (QOL) becomes more closely coupled with independence and hence timely access to rehabilitation is important. The objectives of this paper are (i) to characterize the disability profiles of people with MS admitted to in-patient rehabilitation in Canada; and (ii) to estimate the extent to which disability profiles differ from admission to discharge.

**Methods:** A longitudinal study of a rehabilitation admission-to-discharge cohort of 3500 people with MS was conducting using latent class analysis on the five Functional Independence Measure (FIM) subscales for Self-care, Transfers, Locomotion, Bladder/Bowel, and Cognition. The extent to which age, sex, and calendar time was associated with latent class membership, at both admission and discharge, was estimated using ordinal logistic regression, and proportional odds model was calculated for each age and sex.

**Results:** A five-class model fit the data at admission and a four-class model fit the data at discharge determined using likelihood ratio G^2^, Akaike’s Information Criterion, and Bayesian Information Criterion.

**Conclusion:** At admission, the disability profiles showed a hierarchical progression across the FIM subscales. The least disabled profile was characterized by locomotion dependency only; the most disabled profile was characterized by dependencies in all subscales except cognition. At discharge, the least disabled class, representing 28% of discharges, was characterized by no dependencies; the most disabled class remained with dependencies (23%) in all areas. The study highlights the importance of reserving scarce rehabilitation services to those with more disability.

## Introduction

Multiple Sclerosis (MS) is a progressive and chronic neurodegenerative condition affecting young adults during the peak productive years of their lives with women affected twice as often as men in North America [1, 2]. Current guidelines for management of MS involves a combination of disease-modifying therapy with wellness efforts and lifestyle modifications such like smoking cessation, weight reduction, and regular physical activity [3]. Studies have shown exercise to be neuroprotective and immunomodulatory [4-11]. The MS population is unique in the context of rehabilitation in that the population is young, mainly women, and the disability has an insidious onset and progresses over time, unlike stroke or trauma, the most common reasons for admissions to in-patient rehabilitation. The most common type of MS is associated with exacerbations and remissions but if the exacerbation is severe and the person is hospitalized, the return to functional level may require rehabilitation services.

As part of an international initiative launched by the European organization, Rehabilitation in Multiple Sclerosis [12], there is interest in understanding variability across countries in rehabilitation services and outcomes for people with MS. Inpatient rehabilitation is an important part of the continuum of rehabilitation care for people with MS. Rehabilitation in MS is directed at maintaining functional independence and autonomy. In France, public health covers access to adequate healthcare, with coordinated, continuous, and interdisciplinary rehabilitation services [13]. For people with MS, interdisciplinary services are offered at the time of diagnosis with access to rehabilitation regardless of age, type of MS, or disability level [13]. This is unfortunately not a case in Canada, and it not known what qualifies a person with MS to be admitted to in-patient rehabilitation. While there is a push to provide rehabilitation services to people with MS, the processes and outcomes are unknown. A newly funded initiative, MSCAN Rehab, is a network of rehabilitation researchers who are dedicated to generating and mobilizing rehabilitation evidence to improve the lives people affected by MS.

Despite a lack of consistency in access to rehabilitation services in Canada, inpatient centers do admit people with MS. A recent paper on rehabilitation admission for young adults with MS (16 to 35 years) showed that while younger people were admitted with a more severe disability profile, they were more likely to improve over the course of rehabilitation [14]. This analysis was made possible because of the National Rehabilitation Reporting System (NRS) collects data from adult inpatient rehabilitation facilities and programs across Canada.

An individual’s function is assessed using the Functional Independence Measure (FIM) which has evidence for validity, reliability, responsiveness, and feasibility in multiple rehabilitation populations when administered by a trained assessor [15-18]. Understanding the disability profiles at admission and discharge will help identify ways to best allocate these scarce resources. Therefore, the objectives of this study are two-fold: (i) to characterize the disability profiles of people with MS admitted to in-patient rehabilitation in Canada; and (ii) to estimate the extent to which disability profiles differ from admission to discharge.

## Methods

### Measurements

The disability profiles were derived from the subscales of the FIM which is mandated for use in Canadian in-patient rehabilitation centers. The FIM was developed in 1997 at the Uniform Data System for Medical Rehabilitation [19, 20]. The instrument has 18-items each scored on 7-point ordinal scale yielding a total score from 18 to 126. The 18 items can also be used to derive six subscale scores with a differing number of items per subscale: Self-care, six items, score range 6 to 42; Transfer, 3 items, score range 3 to 21; Locomotion, 2 items, score range 2 to 14; Sphincter control (Bladder/Bowel), 2 items, score range 2 to 14; and Social cognition, 3 items, score range 3 to 21. The Communication subscale was excluded as communication impairments are not a feature of MS [21, 22]. Additional variables were age, sex, and situational variables related to living arrangement, work status, and discharge services. Table 1 shows how the scoring (1 to 7) for items for each of the seven FIM subscales was converted to levels of dependency to define disability profiles.

**Table 1:** Item scoring for each FIM subscale and dependency levels used in the latent class analysis

| Score | Description | Dependency Level | Description |
| --- | --- | --- | --- |
| 1 | Total assistance; | 1 | Moderate to total assistance |
| 2 | Maximal assistance |  |  |
| 3 | Moderate assistance |  |  |
| 4 | Minimal assistance | 2 | Minimal assistance |
| 5 | Supervision | 3 | Supervision |
| 6 | Modified independence | 4 | Independent with or without an aid |
| 7 | Complete independence |  |  |

### Design

A longitudinal study of a rehabilitation admission-to-discharge cohort of people with MS (age: 16 to 100 years) was conducted.

### Source

Data was obtained from the Canadian Institute of Health Information (CIHI) NRS from January 1, 2000, to June 30, 2018. The NRS uses the World Health Organization’s International Classification of Disease (ICD-10-CA code G35) for diagnosis of Multiple Sclerosis [23]. The NRS is a data repository of adult inpatient rehabilitation facilities and programs across nine provinces in Canada (except Quebec). The different facilities and programs that contribute data to the NRS include specialized facilities, hospital rehabilitation units and programs, and designated rehabilitation beds. The data is collected at two time points, on admission and at discharge, and submitted NRS online at each quarter (https://www.cihi.ca). Anonymized data was obtained under the Graduate Student Data Access Program to authors Mate KKV (graduate student), and supervisors Mayo N, and Hum S. Previous analysis on this dataset restricted the sample to people from 16 to 35 years of age (n=457). Therefore, this analysis includes the sample reported on previously [24].

### Analysis

Distributional parameters were calculated for each of the seven age categories (16-20, 21-30, 31- 40, 41-50, 61-70, and ≥71 years) for sex, situational variables at admission, discharge, and change in-between (communication subscale excluded). FIM total scores at admission and discharge were graphically presented across age categories and linear regression was used to estimate the association between age category and these values (adjusted for sex). FIM efficiency was calculated as a difference between discharge and admission total FIM scores and length of stay (discharge - admission FIM total score/length of stay).

Latent class analysis was conducted separately at admission and discharge using the five FIM subscales for **<u>S</u>**elf-care, **<u>T</u>**ransfers, **<u>L</u>**ocomotion, **<u>B</u>**ladder/Bowel, and **<u>C</u>**ognition, at admission and discharge [25]. The LCA is an iterative process that finds the best fitting class for each person. The LCA yields posterior probabilities for each person for each class which can be used to assign people deterministically to a class based on their highest probabilities. In addition to the posterior probabilities, good fit of the model to the data was determined using likelihood ratio G^2^, Akaike’s Information Criterion (AIC), and Bayesian Information Criterion [26]. Each person was assigned to a latent class at admission and at discharge and the cross-tabulation of these classes was presented. The probability of people in a given admission class transitioning into a particular discharge class was calculated.

The extent to which age, sex, and calendar time was associated with latent class membership, at both admission and discharge, was estimated using ordinal logistic regression, the proportional odds model, as the latent classes showed a natural ordering from most disabled to least disabled. Proportional odds ratios (PORs), along with 95% confidence intervals (95% CI), were calculated for each age category in comparison to the oldest category (referent: age ≥71 years). For sex, the referent category was men, and for calendar time the referent category was year 2018. After visual inspection of the data, calendar time was treated linearly. PORs >1 indicate that people in the strata show an increase odd of being in a less disabled class at admission and at discharge. Data analysis was conducted using the Statistical Analysis System (SAS)^TM^ version 9.4.

## Results

Table 2 presents the demographic and situational characteristics of the MS population undergoing inpatient physical rehabilitation according to age category (n=3500). The mean age and SD for each age category is presented. The majority of the sample were women, range 64% to 74% across age categories. All other variables showed differences with age. For example, 83% of people in the youngest age category were ambulatory versus 53% in the oldest age category. Similarly, length of stay ranged from 26.5 days to a peak of 42.2 days for age category 51 to 60 years and 37.5 days for the oldest age group. At discharge, 64% in the youngest age group were discharged home without health services compared with only 10.5% in the oldest age.

**Table 2:**
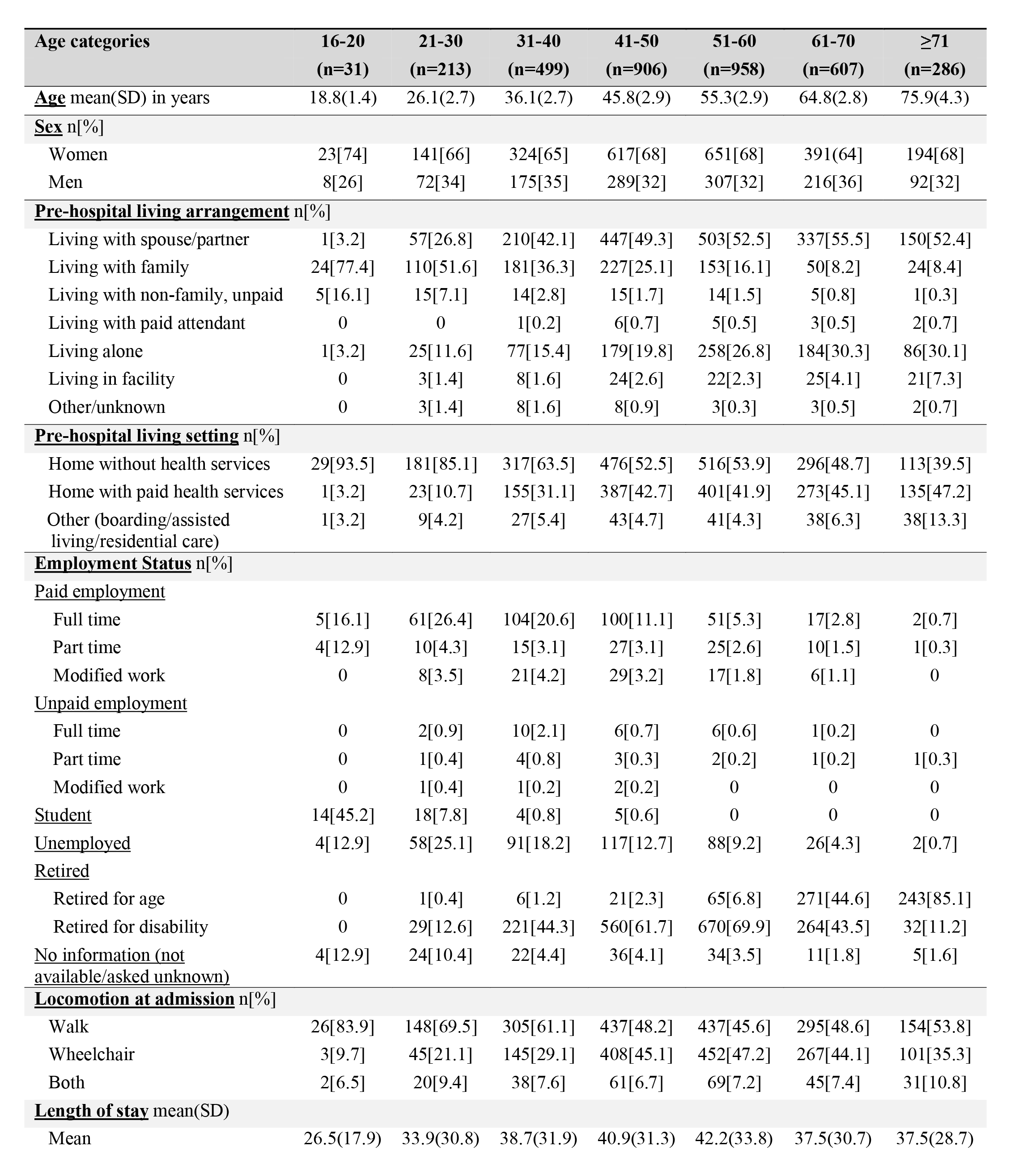
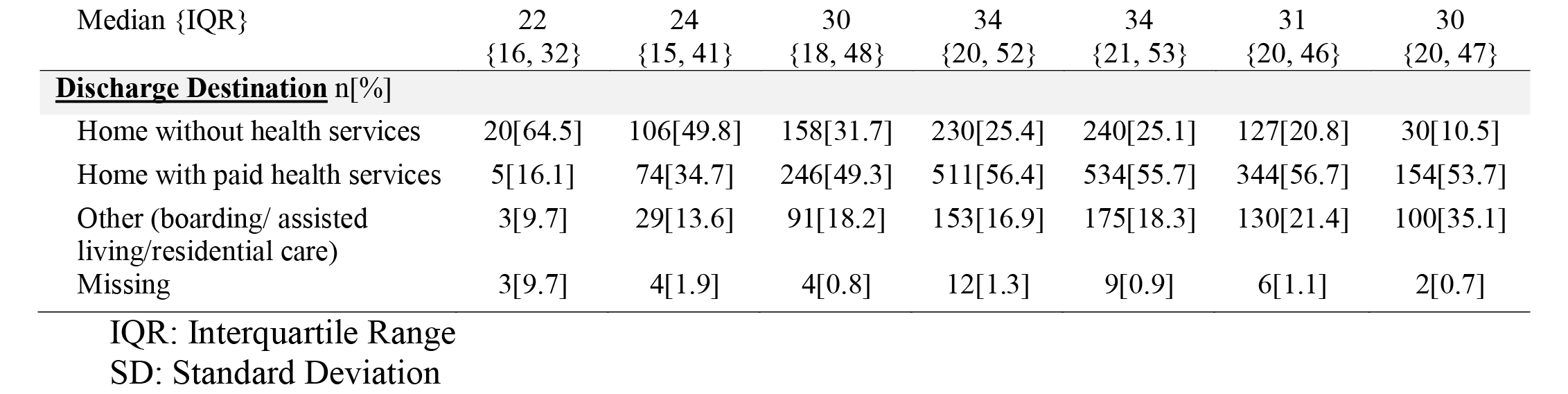
Demographic Characteristics of the Population (n=3500)

| **Age categories** | **16-20**<br>**(n=31)** | **21-30**<br>**(n=213)** | **31-40**<br>**(n=499)** | **41-50**<br>**(n=906)** | **51-60**<br>**(n=958)** | **61-70**<br>**(n=607)** | **≥71**<br>**(n=286)** |
| --- | --- | --- | --- | --- | --- | --- | --- |
| **Age** mean(SD) in years | 18.8(1.4) | 26.1(2.7) | 36.1(2.7) | 45.8(2.9) | 55.3(2.9) | 64.8(2.8) | 75.9(4.3) |
| **Sex** n[%] |  |  |  |  |  |  |  |
| Women | 23[74] | 141[66] | 324[65] | 617[68] | 651[68] | 391[64] | 194[68] |
| Men | 8[26] | 72[34] | 175[35] | 289[32] | 307[32] | 216[36] | 92[32] |
| **Pre-hospital living arrangement** n[%] |  |  |  |  |  |  |  |
| Living with spouse/partner | 1[3.2] | 57[26.8] | 210[42.1] | 447[49.3] | 503[52.5] | 337[55.5] | 150[52.4] |
| Living with family | 24[77.4] | 110[51.6] | 181[36.3] | 227[25.1] | 153[16.1] | 50[8.2] | 24[8.4] |
| Living with non-family, unpaid | 5[16.1] | 15[7.1] | 14[2.8] | 15[1.7] | 14[1.5] | 5[0.8] | 1[0.3] |
| Living with paid attendant | 0 | 0 | 1[0.2] | 6[0.7] | 5[0.5] | 3[0.5] | 2[0.7] |
| Living alone | 1[3.2] | 25[11.6] | 77[15.4] | 179[19.8] | 258[26.8] | 184[30.3] | 86[30.1] |
| Living in facility | 0 | 3[1.4] | 8[1.6] | 24[2.6] | 22[2.3] | 25[4.1] | 21[7.3] |
| Other/unknown | 0 | 3[1.4] | 8[1.6] | 8[0.9] | 3[0.3] | 3[0.5] | 2[0.7] |
| **Pre-hospital living setting** n[%] |  |  |  |  |  |  |  |
| Home without health services | 29[93.5] | 181[85.1] | 317[63.5] | 476[52.5] | 516[53.9] | 296[48.7] | 113[39.5] |
| Home with paid health services | 1[3.2] | 23[10.7] | 155[31.1] | 387[42.7] | 401[41.9] | 273[45.1] | 135[47.2] |
| Other (boarding/assisted living/residential care) | 1[3.2] | 9[4.2] | 27[5.4] | 43[4.7] | 41[4.3] | 38[6.3] | 38[13.3] |
| **Employment Status** n[%] |  |  |  |  |  |  |  |
| **<u>Paid employment</u>** |  |  |  |  |  |  |  |
| Full time | 5[16.1] | 61[26.4] | 104[20.6] | 100[11.1] | 51[5.3] | 17[2.8] | 2[0.7] |
| Part time | 4[12.9] | 10[4.3] | 15[3.1] | 27[3.1] | 25[2.6] | 10[1.5] | 1[0.3] |
| Modified work | 0 | 8[3.5] | 21[4.2] | 29[3.2] | 17[1.8] | 6[1.1] | 0 |
| **<u>Unpaid employment</u>** |  |  |  |  |  |  |  |
| Full time | 0 | 2[0.9] | 10[2.1] | 6[0.7] | 6[0.6] | 1[0.2] | 0 |
| Part time | 0 | 1[0.4] | 4[0.8] | 3[0.3] | 2[0.2] | 1[0.2] | 1[0.3] |
| Modified work | 0 | 1[0.4] | 1[0.2] | 2[0.2] | 0 | 0 | 0 |
| **Student** | 14[45.2] | 18[7.8] | 4[0.8] | 5[0.6] | 0 | 0 | 0 |
| **Unemployed** | 4[12.9] | 58[25.1] | 91[18.2] | 117[12.7] | 88[9.2] | 26[4.3] | 2[0.7] |
| **Retired** |  |  |  |  |  |  |  |
| Retired for age | 0 | 1[0.4] | 6[1.2] | 21[2.3] | 65[6.8] | 271[44.6] | 243[85.1] |
| Retired for disability | 0 | 29[12.6] | 221[44.3] | 560[61.7] | 670[69.9] | 264[43.5] | 32[11.2] |
| **No information (not available/asked unknown)** | 4[12.9] | 24[10.4] | 22[4.4] | 36[4.1] | 34[3.5] | 11[1.8] | 5[1.6] |
| **Locomotion at admission** n[%] |  |  |  |  |  |  |  |
| Walk | 26[83.9] | 148[69.5] | 305[61.1] | 437[48.2] | 437[45.6] | 295[48.6] | 154[53.8] |
| Wheelchair | 3[9.7] | 45[21.1] | 145[29.1] | 408[45.1] | 452[47.2] | 267[44.1] | 101[35.3] |
| Both | 2[6.5] | 20[9.4] | 38[7.6] | 61[6.7] | 69[7.2] | 45[7.4] | 31[10.8] |
| **Length of stay** mean(SD) |  |  |  |  |  |  |  |
| Mean | 26.5(17.9) | 33.9(30.8) | 38.7(31.9) | 40.9(31.3) | 42.2(33.8) | 37.5(30.7) | 37.5(28.7) |

| Median {IQR} | 22<br>{16, 32} | 24<br>{15, 41} | 30<br>{18, 48} | 34<br>{20, 52} | 34<br>{21, 53} | 31<br>{20, 46} | 30<br>{20, 47} |
| --- | --- | --- | --- | --- | --- | --- | --- |
| **Discharge Destination** n[%] |  |  |  |  |  |  |  |
| Home without health services | 20[64.5] | 106[49.8] | 158[31.7] | 230[25.4] | 240[25.1] | 127[20.8] | 30[10.5] |
| Home with paid health services | 5[16.1] | 74[34.7] | 246[49.3] | 511[56.4] | 534[55.7] | 344[56.7] | 154[53.7] |
| Other (boarding/ assisted living/residential care) | 3[9.7] | 29[13.6] | 91[18.2] | 153[16.9] | 175[18.3] | 130[21.4] | 100[35.1] |
| Missing | 3[9.7] | 4[1.9] | 4[0.8] | 12[1.3] | 9[0.9] | 6[1.1] | 2[0.7] |

IQR: Interquartile Range

Table 3 presents the mean (SD) of five FIM subscales at admission, discharge, and change scores across seven age groups with lower scores observed with older age groups.

**Table 3:**
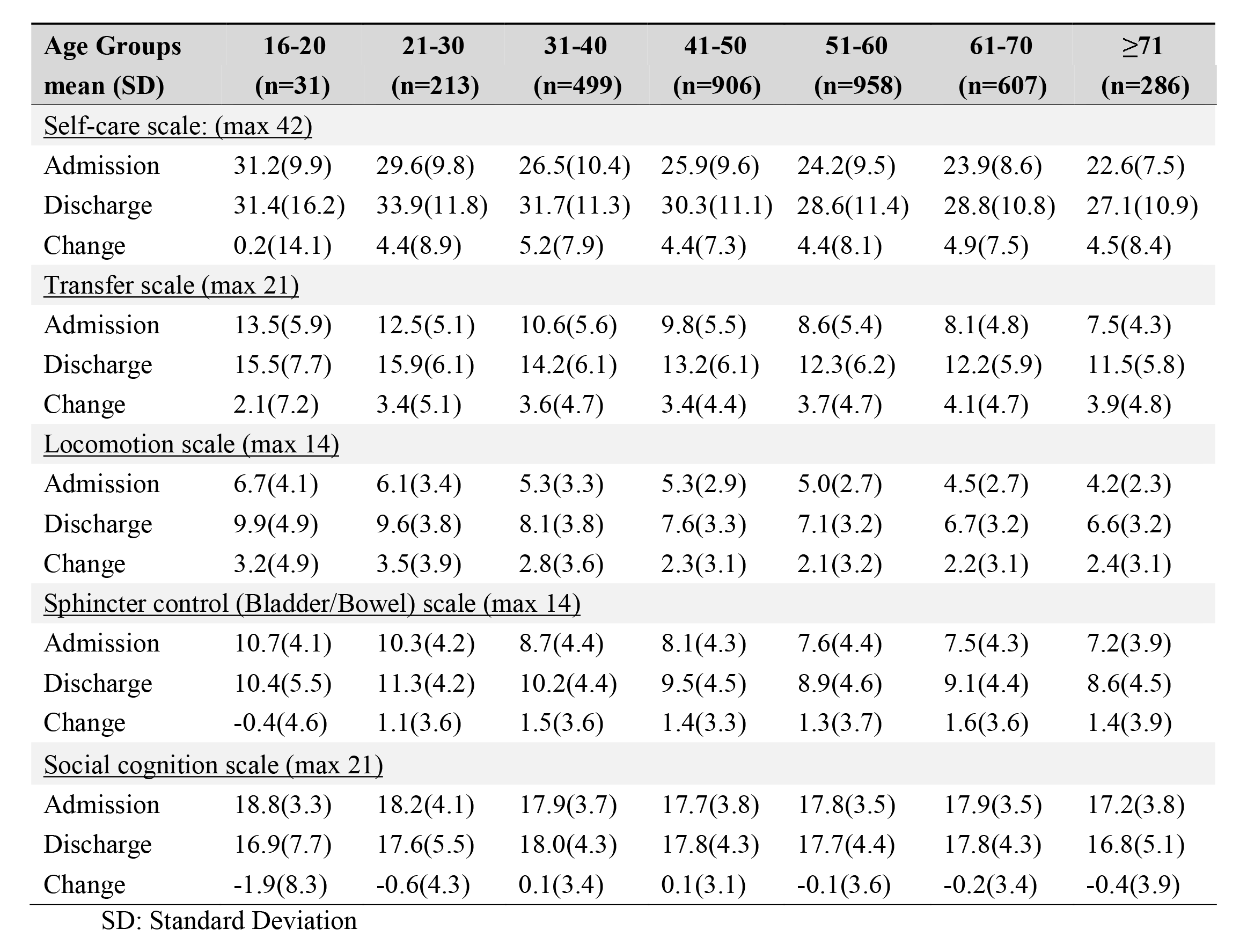
FIM subscales and total scores (mean and SD) at admission and discharge by age groups

Figure 1 depicts total FIM scores and SE at admission and discharge. Overall, the FIM scores at admission and discharge were lower with increasing age category (admission: β: −3.1; SE: 0.2; discharge β: −3.3; SE: 0.3 where β /SE is equivalent to a t-test with the two-sided critical value of 1.95). There was no effect of sex on admission and discharge scores. Also shown is the change score among those with complete data. Change scores were not affected by age category and there was a small but not clinically meaningful difference by sex (β: +1.42; SE: 0.52). When linked to length of stay, change in total FIM scores translates to FIM efficiency (mean change in FIM / days in rehabilitation); in this population the average FIM efficiency across all people admitted was 0.54 (95% CI: 0.47 to 0.53).

**Figure 1:**
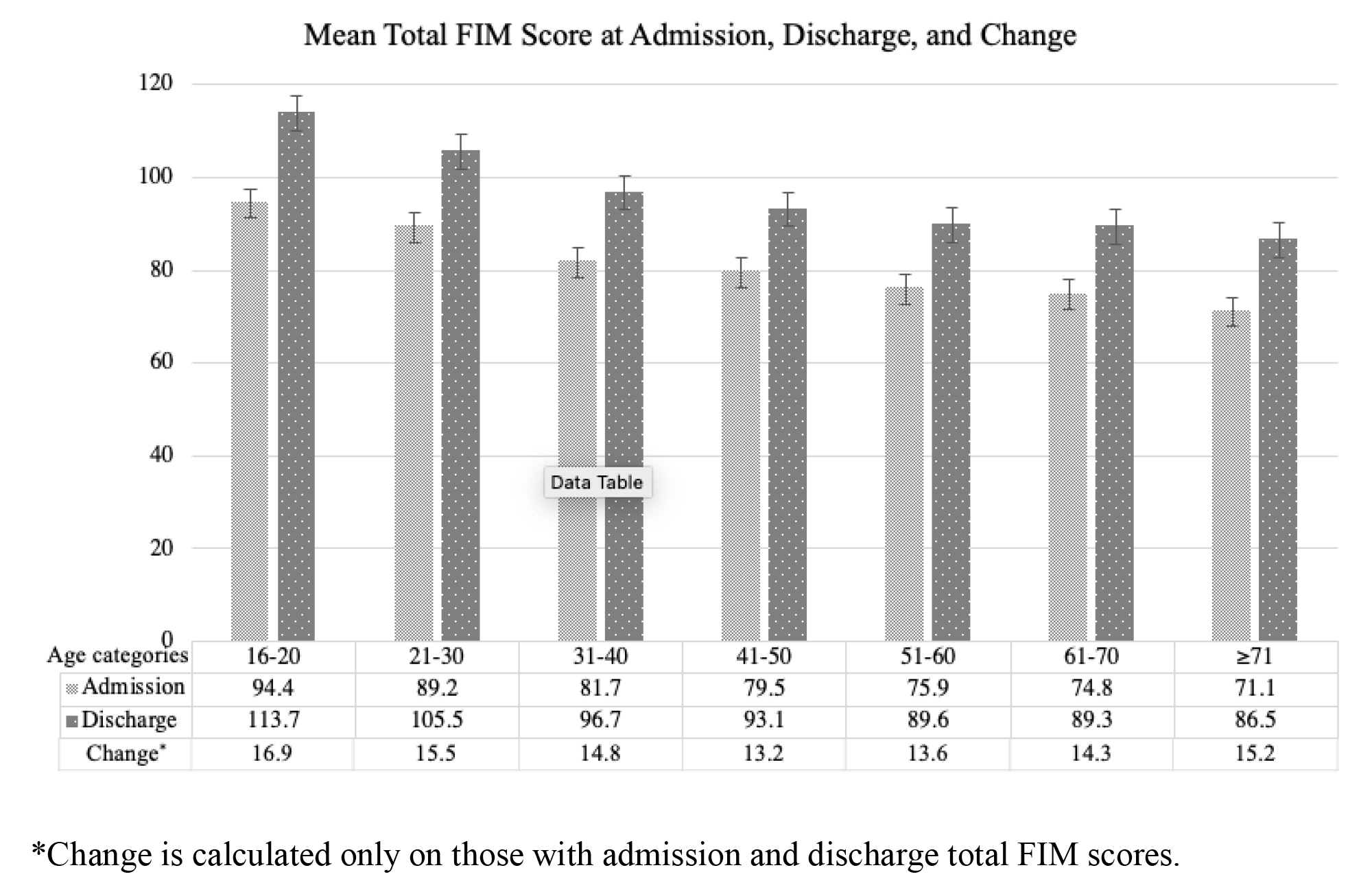
Mean of total FIM score at admission, discharge, and change

Figure 2 shows the class membership at admission on FIM scale category according to areas of dependency (<u>S</u>elf-care, <u>T</u>ransfers, <u>L</u>ocomotion, <u>B</u>ladder, and <u>C</u>ognition). The L_Dependent (Locomotion dependent; 44144; n=523) group at admission was independent in all other subscales except locomotion. The STL_1 class (<u>S</u>elf-care, <u>T</u>ransfers, <u>L</u>ocomotion, <u>B</u>ladder, and <u>C</u>ognition; 3,<3,144; n=465) was less involved (relatively more independent) than STL_2 class (<u>S</u>elf-care, <u>T</u>ransfers, <u>L</u>ocomotion, <u>B</u>ladder, and <u>C</u>ognition; 2,2.5,144; n=208). Both these classes were independent in Bladder and Cognition. Similarly, the class STLB_1 (<u>S</u>elf-care, <u>T</u>ransfers, <u>L</u>ocomotion, <u>B</u>ladder, and <u>C</u>ognition; 21114; n=1099) was better than STLB_2 class (<u>S</u>elf-care, <u>T</u>ransfers, <u>L</u>ocomotion, <u>B</u>ladder, and <u>C</u>ognition; 11114; n=1205). Both these classes were intendent in cognition. The attached table presents, for each latent class (n=5), the average posterior probabilities of the people in each dependence level (1 to 4) for each of the FIM subscales. The latent class is defined according to the highest posterior probabilities within each subscale (shaded areas). For example, L_Dependent comprises people who are highly likely to be most independent in self-care (level 4; PP = 0.8415). Similarly, this latent class comprises people who have highest posterior probability of being independent in transfers (PP=0.6271) but the mostly likely to dependent for locomotion. For bladder and cognition this class comprises people mostly likely to independent in these two functional areas.

**Figure 2:**
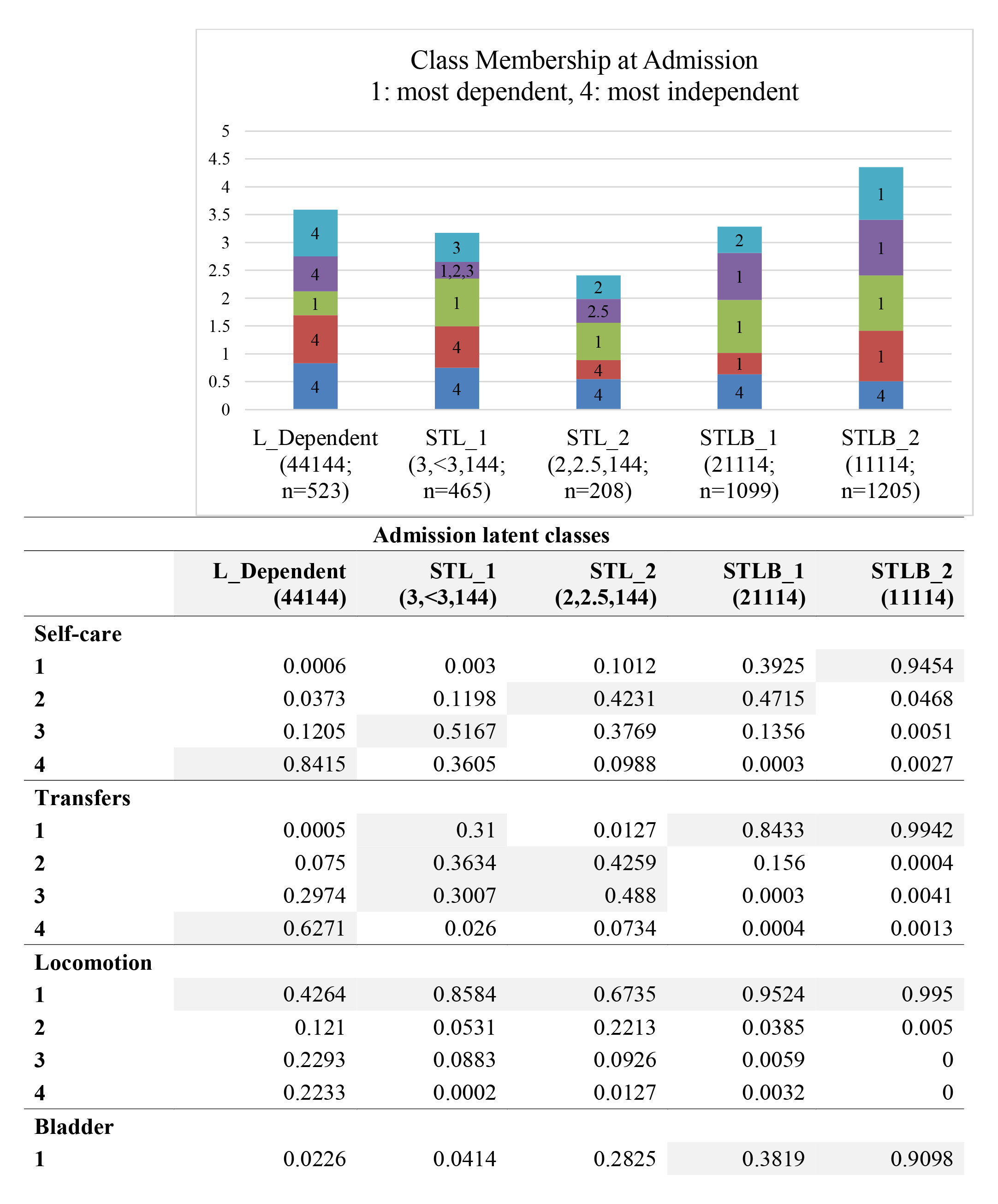
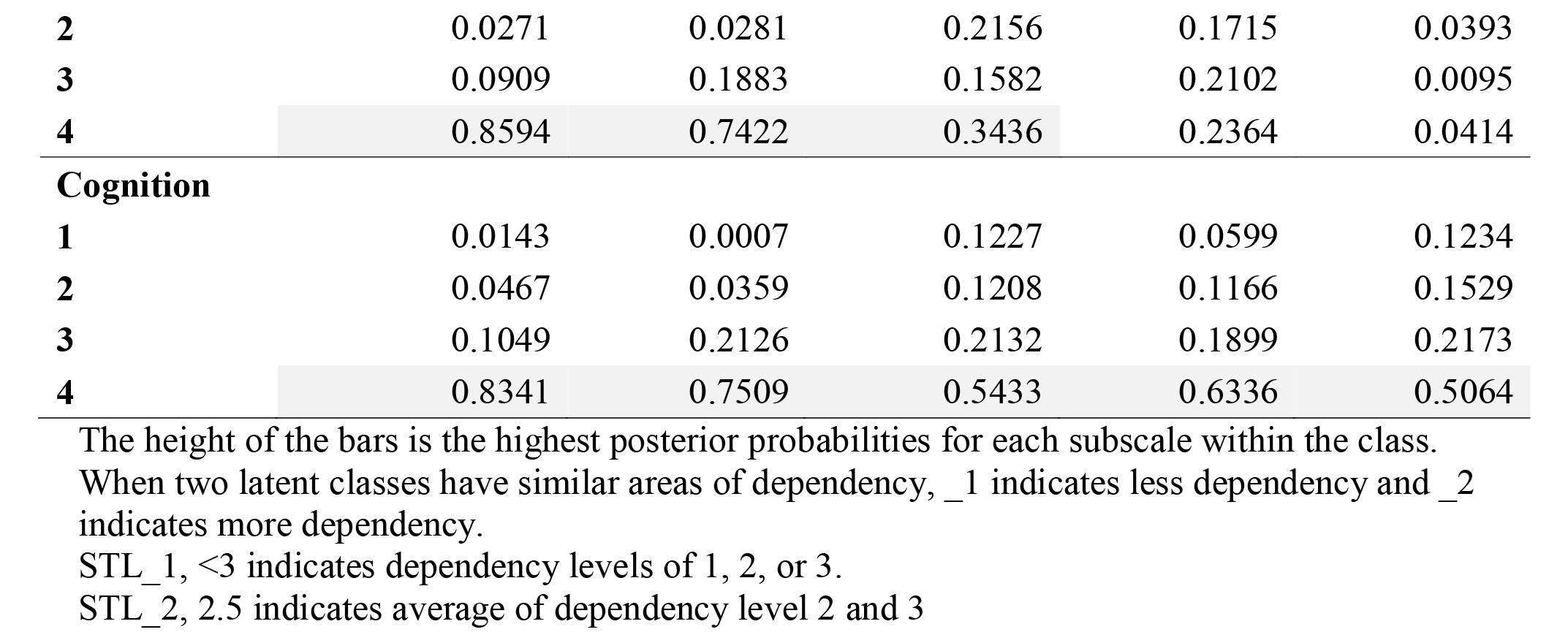
Latent classes based FIM scale category scores at admission labeled according to areas of dependency (**<u>S</u>**elf-care, **<u>T</u>**ransfers, **<u>L</u>**ocomotion, **<u>B</u>**ladder/Bowel, **<u>C</u>**ognition).

Figure 3 presents the class membership at discharge on FIM scale category according to areas of dependency (<u>S</u>elf-care, <u>T</u>ransfers, <u>L</u>ocomotion, <u>B</u>ladder, and <u>C</u>ognition). The ‘None’ class was independent on FIM subscales at discharge (44444; n=946). The TL class (<u>T</u>ransfer and <u>L</u>ocomotion; 43144; n=1010) was dependent on transfer and locomotion subscale of the FIM scale. The STLB_1 (<u>S</u>elf-care, <u>T</u>ransfer, <u>L</u>ocomotion, and <u>B</u>ladder; 22114; n=678), and STLB_2 (<u>S</u>elf-care, <u>T</u>ransfer, <u>L</u>ocomotion, and <u>B</u>ladder; 11114; n=771) was dependent on four subscales to a varying degree with _2 being more dependent than _1. The table presents the posterior probabilities for each of the FIM subscales across the five discharge classes. Shaded grey are the highest posterior probabilities.

**Figure 3:**
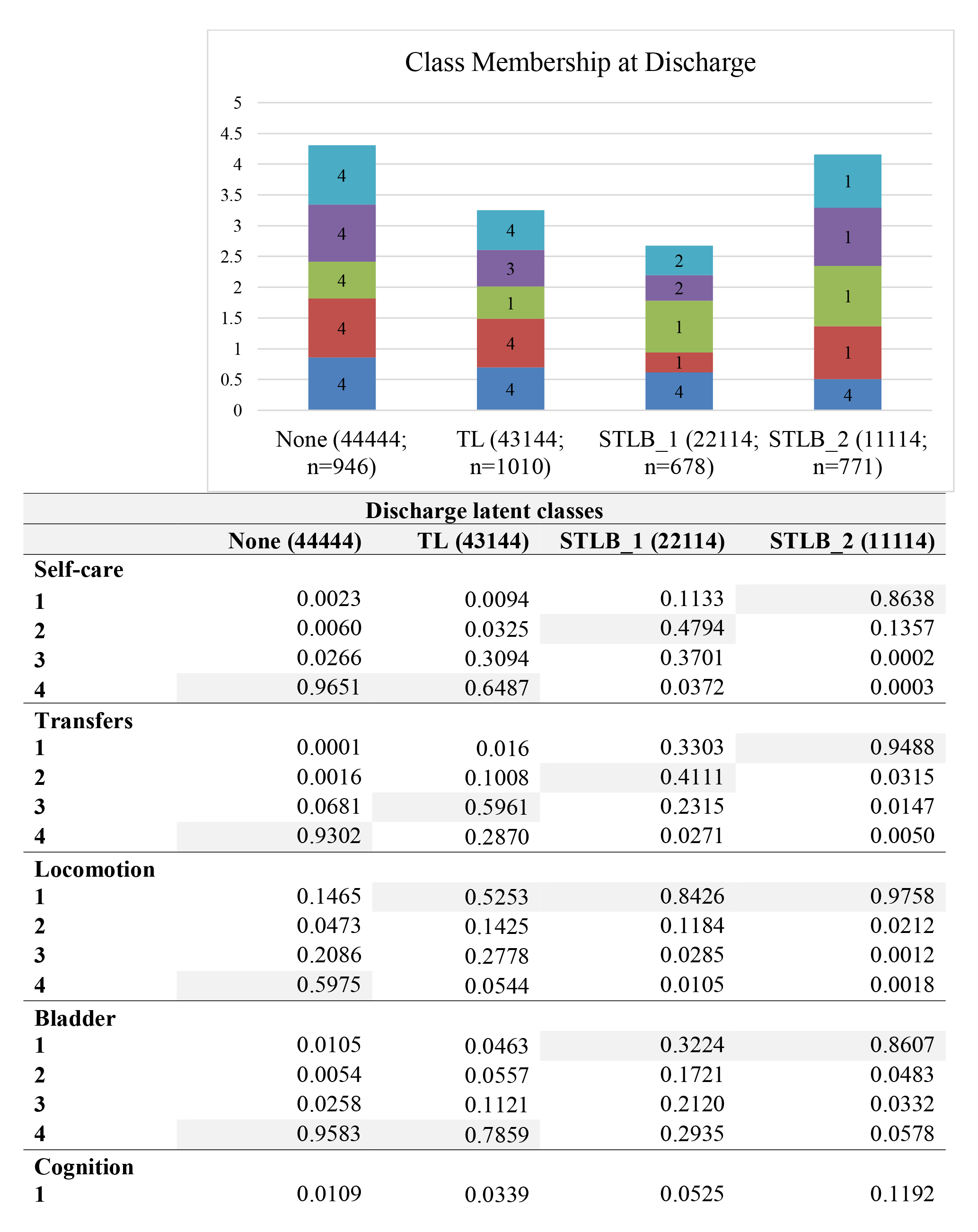
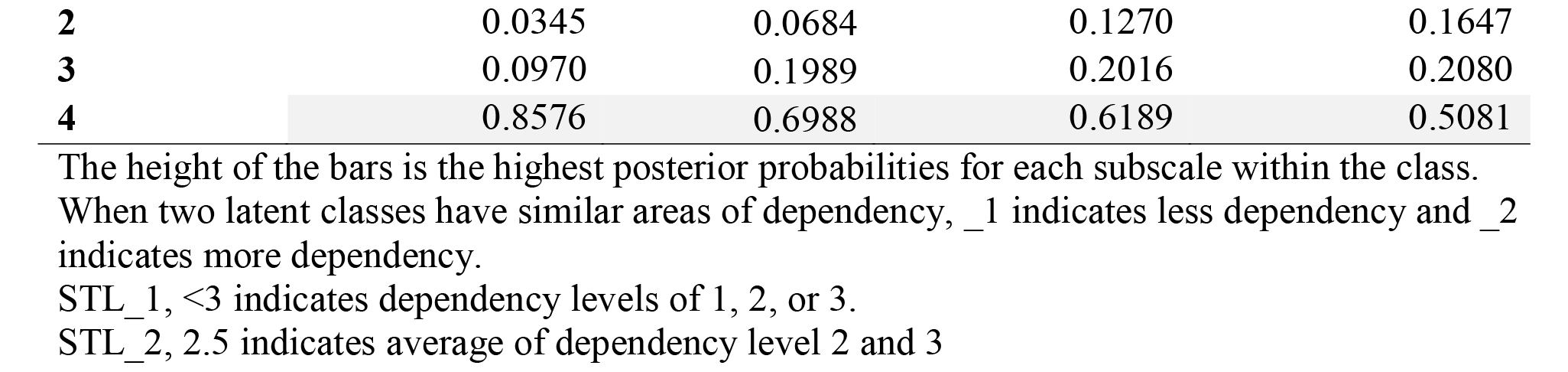
Latent classes based FIM scale category scores at discharge labeled according to areas of dependency (**<u>S</u>**elf-care, **<u>T</u>**ransfers, **<u>L</u>**ocomotion, **<u>B</u>**ladder/Bowel, **<u>C</u>**ognition)

Table 4 shows a five-class model fit the data at admission and a four-class model fit the data at discharge (shaded rows). The latent class model selection is driven statistically by AIC and BIC parameters and theoretically by meaningful interpretation of the different classes. Among the different models at admission, the five-class model was the best fitting with lowest AIC (850.9) and highest BIC (1337.6) values. For discharge, the AIC (930.3) and BIC (1316.6) values were optimal at four-class model level. Clinically both models, five class at admission and a four class at discharge, were meaningful and interpretable.

**Table 4:** Comparison of models of FIM scale scores at admission and discharge

| Number of classes | Likelihood ratio $G^2$ | Degrees of freedom | AIC | BIC |
| --- | --- | --- | --- | --- |
| **Admission** |  |  |  |  |
| 2 | 1607.1 | 992 | 1669.1 | 1860.1 |
| 3 | 887.3 | 976 | 981.3 | 1270.8 |
| 4 | 766.2 | 960 | 892.2 | 1280.4 |
| 5 | 692.9 | 944 | 850.9 | 1337.6 |
| **Discharge** |  |  |  |  |
| 2 | 2284.6 | 992 | 2346.6 | 2536.7 |
| 3 | 887.3 | 976 | 981.3 | 1270.8 |
| 4 | 804.3 | 960 | 930.3 | 1316.6 |
| 5 | 743.4 | 944 | 901.4 | 1385.9 |

Note: Shaded rows indicates selected model.
AIC = Akaike's Information Criterion
BIC = Bayesian Information Criterion

Figure 4 shows the probabilities of transitioning from an admission class to a discharge class with tabulation showing the number of people in each transitional cell used to calculate the transitional probabilities. The X-axis labels the admission classes, and the Y-axis indicates the transitional probabilities. For examples, of the 516 in the L_Dependent class at admission, all but one person transitioned to a lower class at discharge (shown in orange). For the 1155 people in the most dependent class at admission all transitioned to a higher class (shown in green) with the greatest proportion transitioning to a class with no dependency; shown in grey is the probability of remaining in the same class. Figure 4 also provides the number of people in each admission/discharge cell; grey shading shows the cells for people transitioning to a lower dependency level indicating a better discharge class (2120/3405=62.3%).

**Figure 4:**
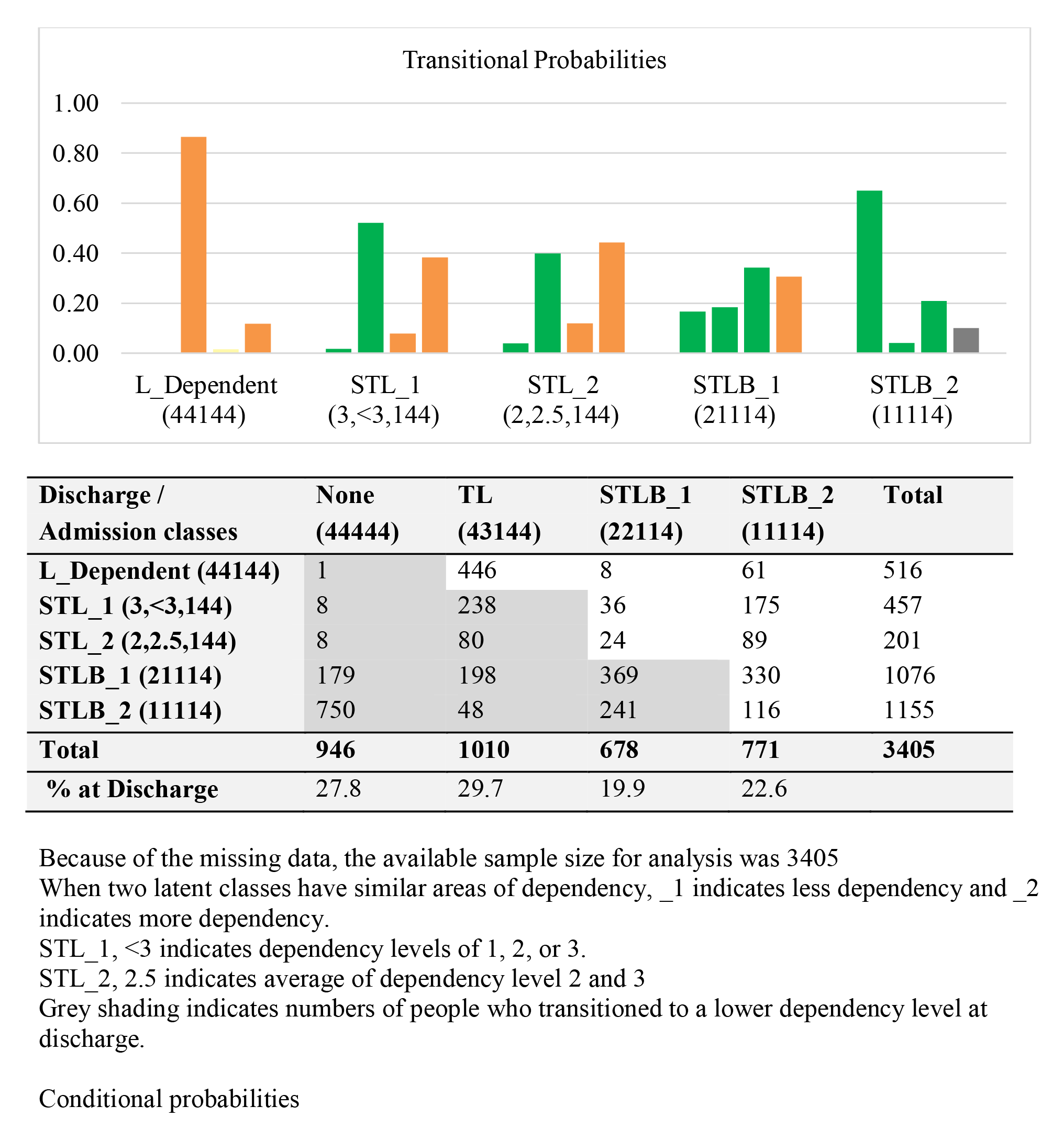
Transitional probabilities and cross-tabulation of FIM latent class membership at admission and discharge labeled according to areas of dependency (**<u>S</u>**elf-care, **<u>T</u>**ransfers, **<u>L</u>**ocomotion, **<u>B</u>**ladder/Bowel, **<u>C</u>**ognition)

Supplemental Table 1 presents the POR and 95% CI across age groups (referent: age ≥71) and sex (referent: men), at admission and discharge. At admission, there was considerable homogeneity in POR across age groups with no strong evidence of differences across age groups. For discharge, there was linear trend of younger people being discharged with less dependency: age 16 to 20, POR 6.7 (95% CI: 3.4 to 13.2); age 61 to 70, POR 1.3 (95% CI: 1.0 to 1.6). There was no association of sex with either admission or discharge class. Supplemental Table 2 shows that there was no pattern of admission class differed over calendar time: POR for linear effect of calendar time was 0.9, 95% CI 0.97 to 0.99.

## Discussion

The present study on 3500 people with MS who underwent in-patient rehabilitation in facilities across Canada (except Quebec) identified five distinct disability profiles at admission (Figure 2) and four at discharge (Figure 3). At admission, the disability profiles showed a hierarchical progression across the FIM subscales of locomotion, transfers, self-care, and bladder/bowel control. The least disabled profile was characterized by locomotion dependency only; the most disabled profile was characterized by dependencies in all subscales except cognition. At discharge, the least disabled class, representing 28% of discharges, was characterized by no dependencies; the most disabled class remained with dependencies (23%) in all areas (except cognition). It is noteworthy that of the people assigned to the least disabled class (locomotor dependency only) at admission, all but one person transitioned to a more dependent class reflecting deterioration (Figure 4). The transitional probabilities, shown in Figure 4, for all other admission classes, showed improvements with those in the most dependent classes showing the greatest probability of transitioning to a better class at discharge.

The only other study using a latent class approach was done by our group on a younger population (16 to 35 years) and concluded that younger people were admitted with a more severe disability profile compared to older young adults but were more likely to benefit from rehabilitation [14]. In contrast, our study did not find that the dependency at admission was associated with age, nor with sex or calendar time (Supplemental Table 1 and 2). However, at discharge there was an effect of age (not sex or calendar time) with younger people more likely to be discharged with lower dependency.

Traditional methods of estimating rehabilitation benefits are often quantified by linking change in total FIM scores to length of stay referred to as FIM efficiency. FIM efficiency from this study was 0.5 (95% CI: 0.47 to 0.53). Other FIM efficiency values from large population-based samples of people with stroke have ranged from 0.48 (95% CI: 0.15, 0.89; n =1995) [27] to 1.59 [19]. The only large sample study of FIM efficiency in people with MS reported a value of 0.23 (95% CI: 0.22, 0.23) over the period 2010 to 2018 [28] considerably lower than ours, although for the same time period. One of the differences between our study and one from UK was the average length of stay was 39 days in Canada and 59 days in the UK. The FIM values at admission and discharge were approximately 6 points lower in the UK study although the change in FIM was similar. Both the Canadian and the UK studies excluded communication subscale from the calculation of total FIM. Communication deficits in people with MS are rare and likely suggestive of other concomitant conditions [19]. From a psychometrics perspective, scores on communication subscale have been shown to not fit well with the overall function construct [29, 30]. And lastly, from clinical point of view, people are less likely to be admitted to rehabilitation units with communication deficits [21, 22].

One explanation for the lower FIM efficiency in the UK is that perhaps people peaked at a lower value but continued in rehabilitation with the hope of achieving greater independence that was not realized. The FIM efficiency values for stroke were logically higher because of natural recovery. FIM efficiencies >0.6 points per day are typically accepted as clinically meaningful, whereas a <0.6 unit/day change represents a poor rate of improvement [31]. In our study, people admitted with the greatest levels of dependency made the most progress (See Supplemental Figure 1). From the policy perspective, this highlights the importance of prioritizing scarce rehabilitation services to those with more disability.

The strength of the study lies in the robust method of latent class analysis on largest MS population in Canada from nine different provincial health facilities to present effectiveness of in-patient rehabilitation service on disability profiles.

There are several limitations to this study. The choice of outcome measures was dictated by policy. FIM is mandated as an outcome for in-patient rehabilitation across Canada. Several studies have demonstrated ceiling effects on the FIM measure [32, 33]. Other performance measure of functional capacity such as 6-Minute Walk Test might have yielded different information on the progress particularly for Latent Class L_Dependent (44144) admitted with poor walking. The generalizability of finding is limited to the rest of Canada as Quebec does to contribute to the data repository at the CIHI. Lastly, having additional information on patient MS duration, use of disease modifying drugs, and date of last relapse would have provided further explanation of the results.

## Conclusion

The disability profiles are clinically coherent and confirm that in-patient rehabilitation in Canada is reserved exclusively for the most disabled adults with MS, particularly those unable to walk without assistance. Substantial proportions of people made gains in ADLs, transfers, and locomotion, gains that would directly impact their QOL. These data support allocating rehabilitation services to this population.

## Data Availability

Data is not available

## Statements & Declarations

### Author Contributions

All authors contributed to the study conception and design. Material preparation, and data analysis were performed by Kedar Mate. The first draft of the manuscript was written by Kedar Mate and Nancy Mayo and all authors commented on previous versions of the manuscript. All authors read and approved the final manuscript.

### Ethics approval

Ethics approval was not required for this project.

---

**Supplemental Table 1:** The relationship of age categories and sex with ordered latent classes at admission and at discharge.

| **Age groups (n/%)** | **POR (95% CI)** | **POR (95% CI)** |
| --- | --- | --- |
|  | **Admission** | **Discharge** |
| 16-20 (31/0.9) | 1.5 (0.7 to 3.0) | <u>6.7 (3.4 to 13.2)</u> |
| 21-30 (213/6.1) | 1.2 (0.9 to 1.7) | <u>4.7 (3.4 to 6.5)</u> |
| 31-40 (499/14.2) | <u>1.3 (1.0 to 1.6)</u> | <u>2.6 (2.0 to 3.4)</u> |
| 41-50 (906/25.9) | 1.2 (0.9 to 1.5) | <u>2.0 (1.6 to 2.6)</u> |
| 51-60 (958/27.4) | <u>1.4 (1.1 to 1.7)</u> | <u>1.5 (1.1 to 1.9)</u> |
| 61-70 (607/17.3) | 1.0 (0.8 to 1.4) | <u>1.3 (1.0 to 1.6)</u> |
| **≥71 (286/8.2)** | **Referent** | **Referent** |
| Sex: Women (Referent: Men) | 1.0 (0.9 to 1.1) | 0.9 (0.8 to 1.0) |

POR: Odds ratios from proportional odds model with latent classes ordered from least disabled Class I to most disabled Class V (admission) and Class IV (discharge). Values that are underlined indicate a statistical association between that age category and the odds of being assigned to a more disabled latent class.

**Supplemental Table 2:** Proportion of people assigned to each admission latent class according to calendar time (2000 to 2018).

| Calendar time<br>(n) | Admission latent classes |  |  |  |  |
| --- | --- | --- | --- | --- | --- |
|  | L_Dependent<br>(44144) | STL_1<br>(3,<3,144) | STL_2<br>(2,2.5,144) | STLB_1<br>(21114) | STLB_2<br>(11114) |
| **2000 (5)** | 20.0 | 20.0 | 0.0 | 0.0 | 60.0 |
| **2001 (75)** | 18.7 | 21.3 | 10.7 | 17.3 | 32.0 |
| **2002 (111)** | 28.8 | 9.9 | 6.3 | 27.0 | 27.9 |
| **2003 (204)** | 13.7 | 19.6 | 5.9 | 25.5 | 35.3 |
| **2004 (219)** | 16.4 | 11.9 | 7.3 | 25.1 | 39.3 |
| **2005 (209)** | 18.2 | 9.6 | 7.2 | 34.9 | 30.1 |
| **2006 (240)** | 15.8 | 15.0 | 5.4 | 26.3 | 37.5 |
| **2007 (216)** | 16.2 | 13.0 | 6.9 | 30.6 | 33.3 |
| **2008 (225)** | 14.2 | 12.4 | 5.8 | 32.9 | 34.7 |
| **2009 (220)** | 13.2 | 14.1 | 6.4 | 33.6 | 32.7 |
| **2010 (213)** | 15.0 | 13.2 | 4.7 | 31.9 | 35.2 |
| **2011 (227)** | 14.5 | 12.3 | 5.3 | 34.4 | 33.5 |
| **2012 (221)** | 14.5 | 17.2 | 8.1 | 32.1 | 28.1 |
| **2013 (227)** | 13.7 | 7.5 | 3.1 | 35.7 | 40.1 |
| **2014 (227)** | 14.1 | 13.7 | 7.5 | 36.6 | 28.2 |
| **2015 (227)** | 13.2 | 11.9 | 3.1 | 33.9 | 37.9 |
| **2016 (229)** | 10.5 | 16.2 | 3.1 | 34.5 | 35.8 |
| **2017 (165)** | 14.6 | 10.3 | 8.5 | 29.7 | 37.0 |
| **2018 (40)** | 5.0 | 12.5 | 7.5 | 32.5 | 42.5 |
| **Total n (%)** | **523 (14.9%)** | **465 (13.3%)** | **208 (5.9)** | **1099 (31.4)** | **1205 (34.5)** |

POR for linear effect of year: 0.9 and 95% CI 0.97 to 0.99.

**Supplemental Figure 1:**
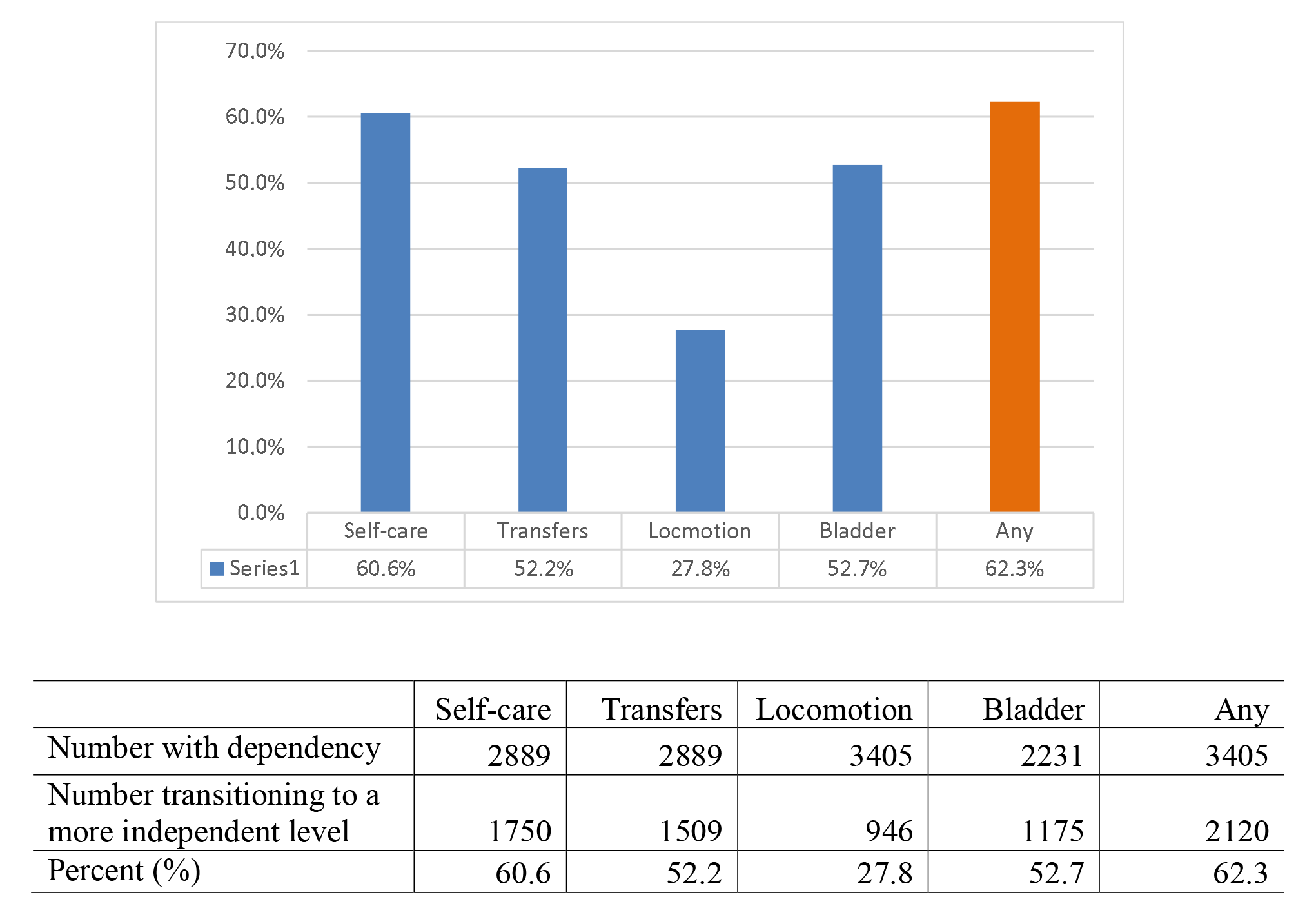
Proportion of people n (%) transitioning to a discharge class with a more independent subscale level

